# Antibodies against the flotillin-1/2 complex in patients with multiple sclerosis

**DOI:** 10.1101/2022.09.14.22278529

**Authors:** Cinta Lleixà, Marta Caballero-Ávila, Elba Pascual-Goñi, Lorena Martín-Aguilar, Nuria Vidal, Clara Tejada, Eduardo Valdés-Hevia, Elisa Zárate, Ana Vesperinas, Roger Collet, Teresa Franco, Laura Martínez-Martínez, Elena Cortés-Vicente, Ricard Rojas-García, Beatriz Gómez-Anson, Anna Gil, Cristina González, Luis Brieva, Sergio Martínez-Yélamos, Luis Querol

## Abstract

Multiple sclerosis (MS) is a tissue-specific autoimmune disease of the central nervous system in which the antigen(s) remains elusive. Antibodies targeting the flotillin-1/2 (FLOT–1/2) complex have been described in 1-2% of the patients in a recent study. Other candidate antigens as anoctamin-2 (ANO2) or neurofascin-155 (NF155) have been previously described in MS patients, although their clinical relevance remains uncertain. Our study aims to analyse the frequency and clinical relevance of antibodies against NF155, ANO2 and the FLOT-1/2 complex in MS.

Serum (n=252) and CSF (n=50) samples from 282 MS patients were included in the study. The control group was composed of 260 serum samples (71 healthy donors and 189 with other neuroinflammatory disorders). Anti-FLOT1/2, anti-ANO2 and anti-NF155 antibodies were tested by cell-based assays using transfected-HEK293 cells. We identified 6 MS patients with antibodies against the FLOT-1/2 complex (2.1%) and 1 MS patient with antibodies against ANO2 (0.35%). All MS patients were negative for anti-NF155 antibodies. Three of the anti-FLOT1/2 positive patients showed anti-FLOT-1/2 positivity in other serum samples extracted at different moments of their disease. IgG subclasses of anti-FLOT-1/2 antibodies were predominantly IgG1 and IgG3.

We confirm that antibodies targeting the Flotillin-1/2 complex are present in a subgroup of patients with MS. Further studies are needed to understand the clinical and pathological relevance of anti-FLOT-1/2 autoantibodies in MS.

## Introduction

Multiple sclerosis (MS) is an autoimmune and neurodegenerative disease of the central nervous system (CNS) that has a heterogeneous clinical presentation and course. The aetiology of MS is likely multifactorial; evidence suggests that various genetic and environmental risk factors contribute to disease manifestation (1). Pathologically, MS is characterized by demyelination, multifocal inflammation, reactive gliosis, and oligodendrocyte and axonal loss, leading to neurological disability (2).

The only validated laboratory biomarker for MS diagnosis is the detection of oligoclonal IgG bands in the CSF (3), that support a prominent role of humoral immunity in this disease (4). Moreover, the epidemiological role of the Epstein-Barr virus (5), the presence of B cells and antibody-secreting plasma cells in CNS demyelinated lesions, and the efficacy of B cell-depleting treatments (6) provide strong evidence for the involvement of B cells and antibodies in MS pathogenesis (7,8).

Despite exhaustive research in autoantibody discovery in MS, the target antigen(s) of the humoral immune response in MS patients remains elusive. A long list of candidate antigens have been found (8), including the glial potassium channel KIR4.1 (9), the flotillin-1/2 complex (FLOT1/2) (10), anoctamin2 (ANO2) (11), or neurofascin-155 (NF155) (12,13), and, most recently antibodies targeting GlialCAM/HEPACAM (14); although most of them have not been replicated in independent cohorts and their clinical relevance remains uncertain (15,16).

Our study aims to analyse the presence of antibodies against NF155, ANO2 and the FLOT-1/2 complex in our MS cohort, and to analyse if there are clinical features associated with these autoantibodies that could suggest that patients harbouring any of these antibodies constitute a differentiated MS subset.

## Materials and methods

### Patients and protocol approvals

Serum (n=252) and CSF (n=50) samples from 282 patients fulfilling the McDonald diagnostic criteria for MS (17) and followed-up in Hospital de la Santa Creu i Sant Pau (n= 188) and Hospital Arnau de Vilanova (n=94) were included in the initial antibody screening. The control group was composed of serum samples from 71 healthy donors and 189 patients with other neuroinflammatory and degenerative disorders: chronic inflammatory demyelinating polyneuropathy (CIDP, n=30), Guillain-Barré syndrome (GBS, n=26), myasthenia gravis (MG, n=44), amyotrophic lateral sclerosis (ALS, n=44), Charcot-Marie-Tooth neuropathy (CMT, n=30), and multifocal motor neuropathy (MMN, n=15) obtained from our Unit biobank (code C.0002365). Serum and CSF samples were obtained, processed and frozen at -80ºC until needed.Additionally, serum samples from 49 MS patients from Hospital de Bellvitge without oligoclonal bands in the CSF were used as confirmation cohort.

Written informed consents were obtained from all subjects for sample handling and data collection. Participation in the study was conducted under a protocol approved by the Institutional Ethics Committee of the Hospital de la Santa Creu i Sant Pau. All methods were carried out in accordance with relevant guidelines and regulations.

### Testing for anti-NF155 and anti-ANO2 antibodies

Anti-NF155 and anti-ANO2 antibodies were tested by cell-based assays using HEK293 transfected cells. Cells were grown in culture dishes with coverslips coated with Poly-D-lysine (Corning, NY, USA), in fetal bovine serum-supplemented DMEM culture medium. After 24 hours, cells were transfected overnight with vectors encoding human NF155 (EX-Z7183-M02, Genecopoeia, Maryland, USA) or ANO2 (RC222601, Origene, Maryland, USA) with Lipofectamine 2000 (Invitrogen, California, US). The day after transfection, cells were fixed with 4 % paraformaldehyde (Santa Cruz Biotechnology, DA, USA) and blocked for 1 hour with 5% goat serum. Double ICC was performed using patients’ sera (1:100) and chicken monoclonal antibody against pan-NF (R&D systems, MI, USA) at 1:1000 or rabbit monoclonal antibody against ANO2 (Origene) at 1:500. Goat anti-human IgG AF594 and goat anti-chicken IgG AF488 (for NF155) or goat anti-rabbit IgG AF488 (for ANO2) (Molecular Probes, Eugene, OR) were used as secondary antibodies at 1:1000 concentration. Finally, culture slides were mounted with Fluoromount medium (Invitrogen) and examined by two independent observers. Images were obtained with an Olympus BX51 Fluorescence Microscope (Olympus Corporation, Tokyo, Japan).

### Testing for anti-FLOT-1/2 antibodies

Anti-flotillin-1/2 antibodies were also tested by cell-based assays using HEK293 cells co-transfected with mammalian-expression vectors encoding human FLOT1 and FLOT2. Our screening study was organized in two steps. First, all the samples were analysed for IgG reactivity on commercial slides containing biochips with cells co-expressing FLOT1 and FLOT2 proteins (Euroimmun, Lübeck, Germany). Each biochip contains one area with transfected cells, and another with non-transfected cells. Slides were incubated with patient’s sera diluted 1:10 in PBS-Tween 0,2% at room temperature for 30 min; followed by incubation with polyclonal goat anti-human IgG FITC (Euroimmun) at room temperature for 30 min. The slides were then mounted with Fluoromount medium and examined by two independent observers. Samples were categorized based on fluorescence intensity of transfected cells in comparison with non-transfected cells and control samples.

Sera from patients showing strong reactivity against the commercial biochips were used for immunocytochemistry over in house co-transfected cells, as explained before. Briefly, mammalian expression vectors encoding human FLOT1 (SC319463, Origene) and FLOT2 (EX-H1820-M02, Genecopoeia) were transfected into HEK293 cells using Lipofectamine 2000. Cells were then fixed with 4% paraformaldehyde and blocked with 5% goat serum. Double ICC was performed using patients’ sera (1:10) and polyclonal rabbit antibodies against FLOT1 (1:100), or FLOT2 (1:250) (Sigma). Goat anti-human IgG AF594 and goat anti-rabbit IgG AF488 (Molecular Probes) were used as secondary antibodies at 1:1000 concentration. To determine the autoantibody subclass, goat anti-human IgG was substituted by mouse anti-human IgG1, IgG2, IgG3 or IgG4 (Southern Biotech).

Finally, culture slides were mounted with Fluoromount medium and examined by two independent observers. Images were obtained with an Olympus BX51 Fluorescence Microscope

### FLOT-1/2 and ANO2 immunoadsorption

Non-transfected, FLOT1-, FLOT2- or FLOT-1/2-cotransfected HEK293 cells (for FLOT-1/2 positive patients) and non-transfected or ANO2-transfected cells (for ANO2 positive patients) were grown in 6-well plates and fixed with 4% paraformaldehyde. Sera showing strong reactivity in immunocytochemistry experiments were diluted in 5% goat serum in PBS, and serially incubated for 1 hour in each of the 6 wells. Then supernatant was collected and its reactivity was tested by immunocytochemistry on FLOT-1/2 co-transfected or ANO2 transfected HEK293 cells, as described above.

### Statistical analysis

Results were analysed by GraphPad Prism v8.0 (GraphPad Software). Statistical comparison of proportions among groups was performed using contingency analysis with the application of a two-tailed Fisher’s exact test, accepting an alpha-level <0.05 for statistical significance.

## RESULTS

### Baseline characteristics

A total of 282 patients fulfilling diagnostic criteria for MS were included in the initial antibody screening (188 from Hospital de la Santa Creu i Sant Pau and 94 from Hospital Arnau de Vilanova). Most patients were women (n=202, 72%) and mean age at diagnosis was 37.19±11.7 years (n=188). Relapsing-remitting multiple sclerosis (RRMS) was the most common subtype of MS (n=239, 85%), 25 patients were diagnosed of primary-progressive MS (PPMS) (9%) and 18 of secondary-progressive MS (SPMS) (6%). CSF study was available in 234 patients and 170 had positive oligoclonal IgG bands (73%).

### Autoantibodies against NF155 and ANO2

We tested 252 sera and 50 CSF samples from 282 patients. None of the patients included in the study had autoantibodies against the NF155 protein. Regarding ANO2 antibodies, we only detected 1 positive serum (1/252, 0.40%). We also tested the CSF and another serum from the same patient but extracted at different moments of the disease, and both samples also showed anti-ANO2 antibodies (**Figure 1**).

**Figure 1.**
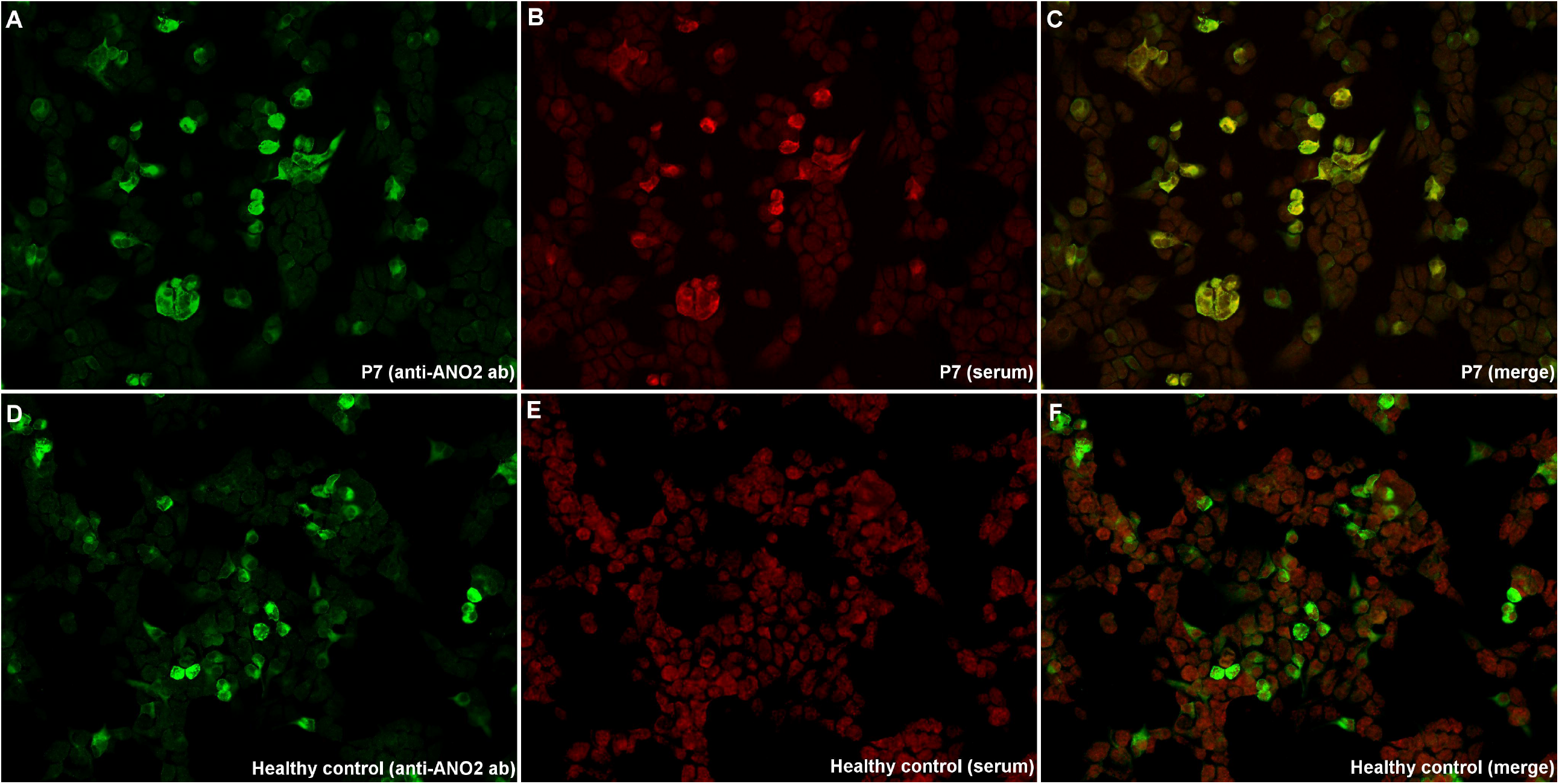
Immunocytochemistry of anti-ANO2 antibodies. HEK293 cells transfected with mammalian-expression vectors encoding human ANO2 using Lipofectamine 2000; double-stained with serum (B, E), and with commercial antibody against ANO2 (A,D). MS patient’s IgG bind to transfected cells (B), and colocalize with ANO2 ab (C); in contrast with the healthy control (E), that does not show any reactivity against ANO2 antibodies.

### Autoantibodies against the FLOT-1/2 complex

We screened the presence of anti-FLOT-1/2 antibodies in 282 patients fulfilling diagnostic criteria for MS (252 sera and 50 CSF), and 260 controls (including healthy donors and patients with other neurological disorders). From the 512 serums tested by immunocytochemistry on commercial biochips containing transfected HEK293 cells, we identified 5 MS patients with antibodies against the flotillin-1/2 complex (5/252, 1.98%), whereas none of the control sera tested showed reactivity in cell-based assays (0/260, 0%) (**Figure 2**). When comparing the results obtained in the two groups, the differences observed were statistically significant (p=0.0283). Moreover, 3 of the anti-FLOT1/2 positive patients showed this positivity in serum samples extracted at different moments of their disease.

**Figure 2.**
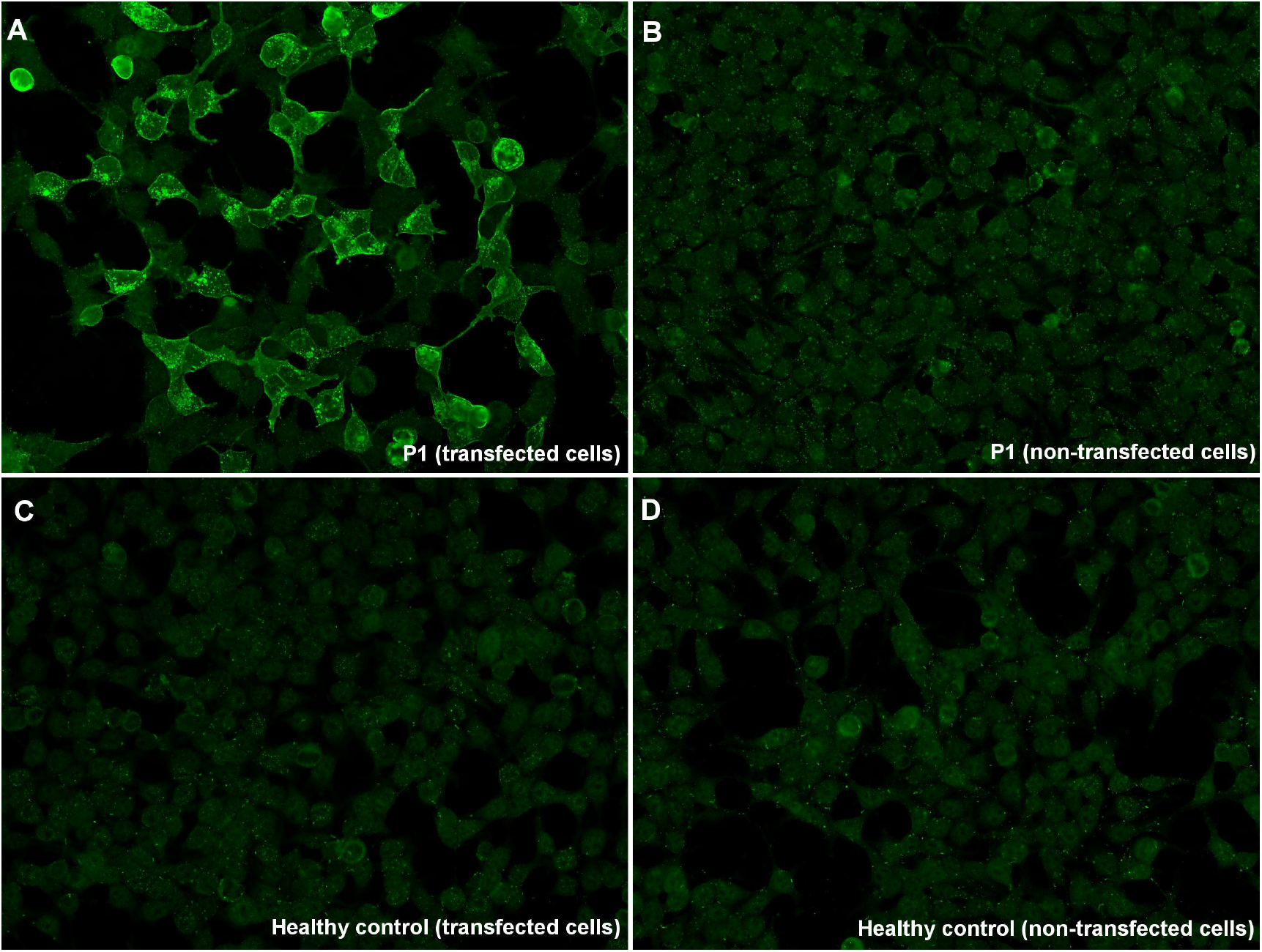
Immunocytochemistry of anti-FLOT1/2 antibodies (commercial slides). Commercial biochips (Euroimmun) containing FLOT-1/2 transfected HEK293 cells (A,C), or non-transfected HEK293 cells (B,D); incubated with serum from MS patient 1 (A,B), and from a healthy donor (C,D). Patient 1 showed strong IgG reactivity against co-transfected cells (A), in comparison with non-transfected cells (B), and with the negative control (C).

Regarding CSF samples, we detected anti-FLOT1/2 antibodies in 1 more MS patient (1/50, 2%). In this case, the autoantibodies were not found in a serum sample of the same patient that was extracted at different moment of the disease.

When assessing the presence of autoantibodies against FLOT-1/2 by immunocytochemistry over *in house* co-transfected HEK293 cells, we confirmed the positivity of the 5 serum and the 1 CSF samples in which we had previously detected anti-FLOT1/2 antibodies with commercial slides (**Figure 3**).

**Figure 3.**
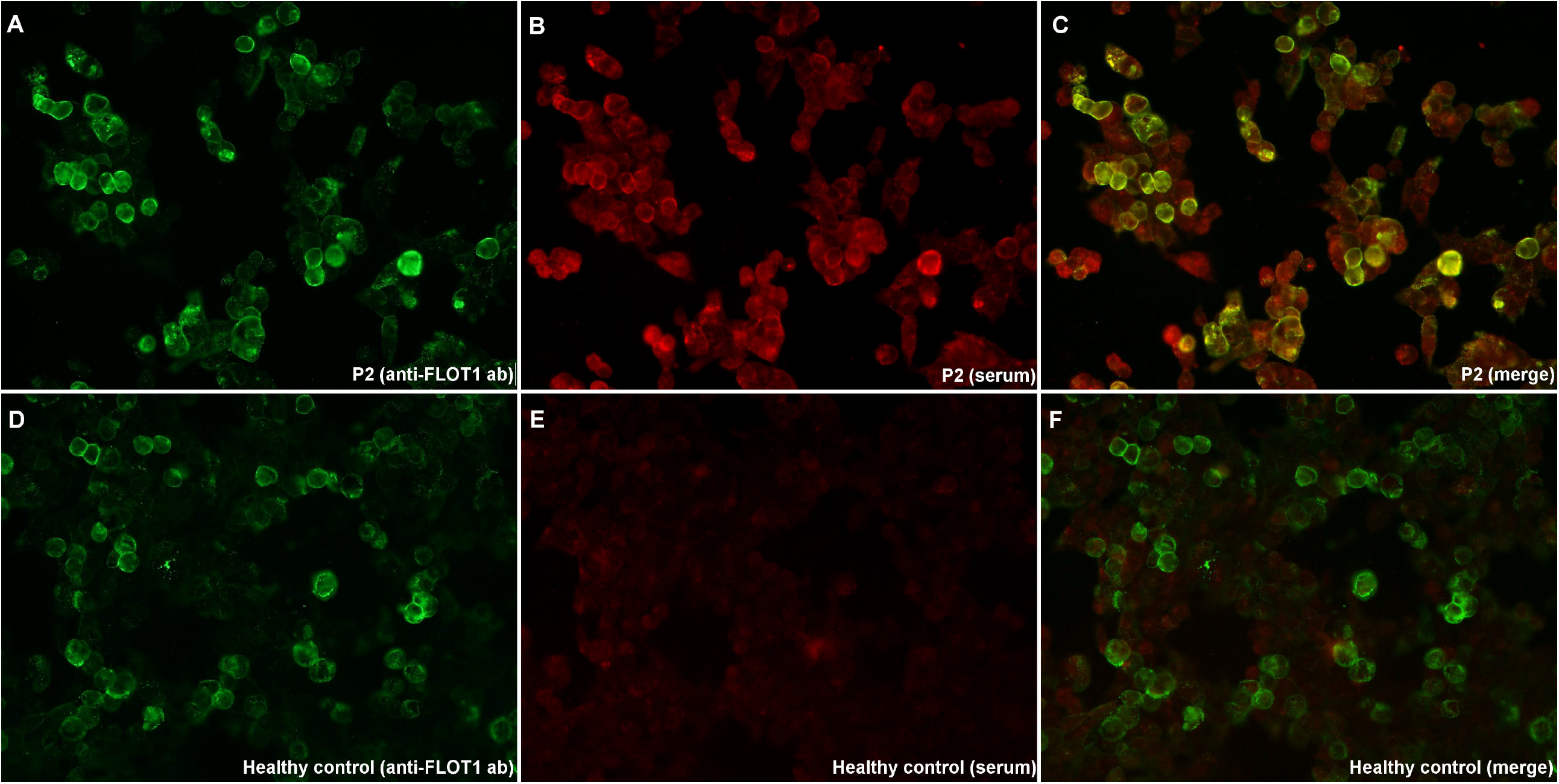
Immunocytochemistry of anti-FLOT1/2 antibodies (*in house* transfection). HEK293 cells co-transfected with mammalian-expression vectors encoding human FLOT1 and FLOT2, using Lipofectamine 2000; double-stained with serum (B, E), and with commercial antibody against FLOT1 (A,D). MS patient’s IgG bind to co-transfected cells (B), and colocalize with FLOT1 ab (C); in contrast with the healthy control (E), that doesn’t show any reactivity against FLOT-1/2 antibodies.

The autoantibody subclass of the positive sera was assessed. The anti-FLOT-1/2 antibodies were exclusively IgG1 in two patients and were consistent in other serum samples extracted at different moment of the disease. Two other patients’ antibodies were IgG3. Finally, in one patient of whom 4 samples were tested, we found that 3 of them had IgG1 anti-FLOT-1/2 and the remaining one had IgG3 antibodies.

We observed that in 3 of 4 patients (75%) that harboured antibodies against the FLOT-1/2 complex, CSF IgG oligoclonal bands were not detected. In the other 2 positive patients oligoclonal IgG bands were not assessed. For that reason, to confirm FLOT-1/2 as a target antigen in MS and a possible correlation between the absence of IgG oligoclonal bands in CSF and the presence of anti-FLOT-1/2 antibodies, we tested these autoantibodies in a cohort from Hospital de Bellvitge including 49 serum samples from patients fulfilling MS criteria in which oligoclonal bands were not detected. However, we did not detect any other patient with antibodies against the FLOT-1/2 complex in this confirmatory cohort.

### Reactivity against FLOT-1/2 and ANO2 antibodies was abrogated after immunoadsorption

We performed immunoadsorption experiments incubating anti-FLOT1/2 positive patients’ sera with non-transfected HEK293 cells, and with HEK cells transfected with FLOT1, FLOT2 or co-transfected with FLOT1/2. Reactivity against the FLOT-1/2 complex was lost after serum pre-adsorption with HEK cells co-expressing FLOT1 and FLOT2, but not after pre-adsorption with cells transfected with FLOT1 or FLOT2 alone, or with non-transfected HEK cells.

We also immunoadsorbed the anti-ANO2 positive patient serum with non-transfected and ANO2-transfected HEK293 cells; and the reactivity against ANO2 was lost after incubation with cells expressing ANO2, but not after incubation with non-transfected cells.

### Clinical features

Clinical and laboratory features of patients with anti-FLOT-1/2 and anti-ANO2 antibodies are summarized in **table 1**. Five out of six anti-FLOT1/2 positive patients fulfilled criteria of RRMS while one patient of PPMS. Four patients were female (67%).

**Table 1.** Clinical and laboratory features of patients with anti-FLOT-1/2 and anti-ANO2 antibodies. RRMS: relapsing-remitting multiple sclerosis; PPMS: primary progressive Multiple Sclerosis; EDSS: Expanded Disability Status Scale; OBC: oligoclonal bands, na= not available.

|  | Anti-FLOT-1/2<br>positive patients |  |  |  |  |  | Anti-ANO2<br>positive patients |
| --- | --- | --- | --- | --- | --- | --- | --- |
|  | P1 | P2 | P3 | P4 | P5 | P6 | P7 |
| **Gender** | Male | Male | Female | Female | Female | Female | Male |
| **Disease duration**<br>**at first sampling**<br>**(years)** | 34 | 6 | 5 | 6 | 13 | 0 | 13 |
| **Optic neuritis** | Yes | No | No | No | No | No | No |
| **Diagnosis** | RRMS | RRMS | RRMS | PPMS | RRMS | RRMS | RRMS |
| **EDSS** | 4 | 4 | 2 | 6.5 | 1 | 1 | 1 |
| **Treatment** | Dymethylfumarate | Natalizumab | Rituximab | Ocrelizumab | Ocrelizumab | None | Fingolimod |
| **Previous**<br>**treatment** | Interferon<br>Laquinimod | Interferon<br>Glatiramer acetate | Interferon<br>Glatiramer acetate<br>Fingolimod<br>Natalizumab | Interferon<br>Teriflunomide | Interferon<br>Glatiramer acetate<br>Teriflunomide<br>Fingolimod | None | Interferon |
| **OCB in CSF** | na | na | Negative | Positive | Negative | Negative | Positive |
| **Number of**<br>**samples with**<br>**Anti-FLOT-1/2** | 1 | 2 | 4 | 1 | 4 | - | - |
| **Serum titer** | 1/1250 | 1/1250 | >1/6250 | 1/50 | >1/6250 | - | - |
| **Serum IgG**<br>**subclass** | IgG3 | IgG1 | IgG1, IgG3 | IgG3 | IgG1 | - | - |
| **CSF titer** | na | na | na | na | na | + | - |

Only one patient had a history of optic neuritis. In two of the three patients treated with anti-CD20 therapies (patient 3 and patient 5), positivity against FLOT-1/2 persisted in serum after starting the treatment (16 months in patient 3 and 2 months in patient 5).

## Discussion

We have identified 6 patients with autoantibodies against the FLOT-1/2 complex and 1 patient with autoantibodies against ANO2 protein in a study to screen for the presence of antibodies against NF155, ANO2 and FLOT1/2, previously described as MS antigens.

In 2017, S. Hahn et al published a report describing the presence of antibodies against the flotillin-1/2 complex in approximately 1-2 % of the MS patients they studied (10). In our cohort, MS patients with anti-FLOT-1/2 autoantibodies in serum are found in 2.1% of the patients.

Despite anti-FLOT1/2 antibodies are very infrequent in MS, we did not detect any positive patient in the control group (that included 260 sera from healthy donors and patients with other neurological disorders). This suggests that anti-FLOT-1/2 antibodies are specific of multiple sclerosis. The presence of these antibodies could define a rare subset of MS patients, as occurs in other neuroinflammatory pathologies; however, despite our work validates the initial description, the clinical and pathophysiological relevance of these autoantibodies in MS remains to be elucidated in autoantibody passive-transfer animal models and larger prospective series are needed to characterize phenotypically anti-FLOT1/2 positive patients.

Data about the presence of flotillin antibodies in other neurological disorders is scarce, with only a few case reports published. A case of limbic encephalitis with positive anti-FLOT-1/2 antibodies in serum and CSF is reported (18). Clinical and radiological findings were compatible with the diagnosis of limbic encephalitis and patient responded to corticosteroids, which supported the autoimmune etiology. In neurodegenerative disorders, a case of atypical dementia associated with anti-FLOT-1/2 antibodies is also published (19), without other similar cases reported. The relevance and pathogenicity of these antibodies in this group of diseases is difficult to elucidate as only isolated cases are reported and further reports would be needed. These reports could also suggest that anti-FLOT1/2 antibodies may arise as a secondary immune response triggered upon brain damage in predisposed individuals. This is the reason why we included controls with other autoimmune (MG, CIDP, MMN, GBS) and degenerative disorders (ALS, CMT).

Flotillin-1 and flotillin-2 are homologous proteins that are found in lipid raft microdomains at the plasma membrane (20). Both flotillins are preferentially associated with each other in hetero-oligomeric complexes (FLOT-1/2) ubiquitously expressed (21). They are involved in axon regeneration and neuronal differentiation (especially in the optic nerve, where these proteins are upregulated), endocytosis, T-lymphocyte activation and membrane protein recruitment (22). One possible explanation for the pathogenicity of autoantibodies against flotillin 1/2 could be that they might impair the recycling of the T-cell receptors (important for activating T-cells)(23), and this failing T-cell activation might lead to cell death causing CNS autoimmunity (24).

In our cohort, no anti-NF155 antibodies were found in the 282 MS patients. The association between the anti-NF155 antibodies and a subgroup of patients with chronic inflammatory demyelinating polyradiculoneuropathy (CIDP) is well established (25) and some patients with combined central and peripheral demyelination are also positive to anti-NF155 antibodies (26). In pure CNS demyelinating disorders such as MS, the proportion of patients with anti-NF155 antibodies differs significantly in different published articles. Anti-NF155 antibodies were detected in 2% of the 243 MS patients tested in the largest study assessing these autoantibodies in MS (12). However, in a previous study, anti-NF155 reactivity was higher than controls in a third of 26 MS patients (27); while in another publication with samples from 20 MS patients the investigators did not find antibodies against NF155 (28). Our report suggests that anti-NF155 antibodies in MS are, at best, infrequent.

The presence of anti-ANO2 antibodies has been anecdotic in our cohort, being present in only one MS patient. Anoctamin-2 is a calcium-activated chloride-channel protein, also named as transmembrane protein 16B which is predominantly expressed in neuronal and muscle tissue (29). Antibodies against ANO2 in MS patients have been widely studied by Ayoglu et al (11)(30). They used a non-conformational antigen array to discover the presence of anti-ANO2 antibodies in the 15.5% of a 1063 MS patients’ cohort. The expression and potential role of anti-ANO2 antibodies in MS remain to be elucidated, but it seems that immunohistochemistry analysis shows a clear increase in ANO2 intensity in the proximity of MS lesions (11).

In conclusion, although we found 6 patients with autoantibodies against the FLOT-1/2 complex and 1 patient with autoantibodies against ANO2 protein, it is not clear if they associate with a specific disease subset or whether these antibodies play a role in MS pathogenesis or represent a nonpathogenic epiphenomenon. For these reasons, further research in larger cohorts is needed to validate these target antigens as biomarkers for MS.

## Data Availability

The datasets used and/or analysed during the current study are available from the corresponding author on reasonable request.

## Acknowledgements

The authors would like to acknowledge the Department of Medicine at the Universitat Autònoma de Barcelona. We also would like to thank all our patients for their patience and collaboration. We want to particularly acknowledge the patients and the Biobank HUB-ICO-IDIBELL (PT17/0015/0024) integrated in the Spanish Biobank Network for their collaboration.

## Author contributions

CL and MCA acquired the data, performed the experiments, analyzed the data and drafted the manuscript; EPG, LMA, NV, CT, EVH, EZ, AV, RC, TF, LMM, ECV, RRG, BGA, AG, CG, LB and SMY acquired samples and data and revised the manuscript for intellectual content; LQ designed and conceptualized the study, interpreted the data and revised the manuscript for intellectual content.

## Competing interests statement

The authors declare that they have no competing interests.

## Notes

### Competing Interest Statement

The authors have declared no competing interest.

### Funding Statement

This work was supported by Roche Spain. MCA was supported by a personal Rio Hortega grant CM21/00101. LMA was supported by a personal Juan Rodes grant JR21/00060.

### Author Declarations

Ethics commitee of Hospital de la Santa Creu i Sant Pau gave ethical approval for this work.

